# Functional annotation of de novo variants found near GWAS loci associated with cleft lip with or without cleft palate

**DOI:** 10.1101/2025.02.21.25322690

**Authors:** Sarah W. Curtis, Laura E. Cook, Kitt Paraiso, Axel Visel, Justin L. Cotney, Jeffrey C. Murray, Terri H. Beaty, Mary L. Marazita, Jenna C. Carlson, Elizabeth J. Leslie-Clarkson

**Affiliations:** Department of Human Genetics, Emory University School of Medicine, Atlanta, GA, 30322; U.S. Department of Energy Joint Genome Institute, Lawrence Berkeley National Laboratory, Berkeley, CA; Environmental Genomics and Systems Biology Division, National Laboratory, Berkeley, California; Department of Surgery, Children’s Hospital of Philadelphia Research Institute, Philadelphia, PA, 19104; Department of Pediatrics, University of Iowa, Iowa City, IA, 52242; Department of Epidemiology, Johns Hopkins Bloomberg School of Public Health, Baltimore, MD, 21205; Center for Craniofacial and Dental Genetics, Department of Oral Biology, University of Pittsburgh, Pittsburgh, PA, 15261; Department of Human Genetics, University of Pittsburgh, Pittsburgh, PA, 15621; Department of Biostatistics and Health Data Science, University of Pittsburgh, Pittsburgh, PA, 15261

## Abstract

Orofacial clefts (OFCs) are the most common craniofacial birth defects, affecting 1 in 700 births, and have a strong genetic basis with a high recurrence risk within families. While many of the previous studies have associated common, noncoding genetic loci with OFCs, previous studies on *de novo* variants (DNVs) in OFC cases have focused on coding variants that could have a functional impact on protein structure, and the contribution of noncoding DNVs to the formation of OFCs has largely been ignored and is not well understood. Therefore, we reanalyzed an existing dataset of DNVs from 1,409 trios with OFCs that had undergone targeted sequencing of known OFC-associated loci. We then annotated these DNVs with information from datasets of predicted epigenetic function during human craniofacial development. Of the 66 DNVs in this cohort, 17 (25.7%) were within a predicted enhancer or promotor region. Two DNVs fell within the same enhancer region (hs1617), which is more than expected by chance (p = 0.0017). The sequence changes caused by these DNVs are predicted to create binding sites not seen in the reference sequence for transcription factors PAX6 and ZBTB7A and to disrupt binding sites for STAT1 and STAT3. This enhancer region is within the same topologically-associated domain as *HHAT*, *SERTAD4*, and *IRF6*, all of which are involved in craniofacial development. All three genes are highly expressed in human neural crest cells. Knockout mice for *Hhat* and *Irf6* have abnormal embryonic development including a cleft palate, and variants in and around *IRF6* are associated with nonsyndromic and syndromic forms of OFCs in humans. Taken together, this suggests that noncoding DNVs contribute to the genetic architecture of OFCs, with a burden of DNVs in OFC trios in enhancer regions near known OFC-associated genes. Overall, this adds to our understanding of the genetic mechanisms that underly OFC formation.

## Introduction

Orofacial clefts (OFCs) are a common birth defect that affect 1 in 700 live births and encompass clefts of the lip (CL), clefts of the palate (CP), and clefts of the lip and palate (CLP) ^1–6^. A majority of OFCs (∼50-70%) are considered to have complex and multifactorial etiology arising from both environmental and genetic risk factors ^7–14^. The genetic architecture of OFCs has been extensively studied. Evidence for a strong genetic component to OFCs comes, in part, from a higher concordance rate in twin studies ^15^ and a 32-fold recurrence risk in first degree relatives of a person with any form of CL ^16^. Several specific genetic loci are also associated with OFCs, with multiple studies having identified common variants near *IRF6*, 8q24, and *FOXE1* ^17–26^, and other studies finding that both coding and regulatory rare variants are associated with OFCs ^27–30^.

While both common and rare variants in non-coding regions have been associated with OFCs, the ability to connect these non-coding regions to the mechanisms through which they lead to OFCs is difficult. One way that a non-coding variant can cause a phenotypic change is by interrupting or interfering with gene regulation ^31^. For example, genetic variation in enhancer or promotor regions could affect how transcription factors bind and thus change how a nearby gene is expressed. However, the genomic regions acting as enhancers are typically cell-type and time-point specific, and it can also be difficult to match enhancers with their target genes, as some enhancers act on the nearest gene and others act on genes several megabases away^32–34^.

OFCs arise due to failure of some aspect of craniofacial development in the first 4-6 weeks of gestation so obtaining the relevant tissue is challenging. Thus, the generation of genomic data from primary human embryonic tissues has lagged behind other, more accessible tissues which has perpetuated the challenges of interpreting non-coding genetic variation in this context. However, there have recently been significant advances in cataloging and predicting regulatory regions involved in human craniofacial development^35^, allowing genome-scale research on role of regulatory genetic variants in individuals with OFCs. Therefore, we decided to address this question by re-annotating *de novo variant*s (DNVs) in people with OFCs and to test if any regulatory regions had a burden of variants. We then used molecular and *in silico* tools to assess the downstream consequences of variants in these regulatory regions.

## Methods

### Study population

This study used data collected from a previous study ^30^, which included 1,409 case-parent trios (4,227 individuals) of Asian or European ancestry recruited from Europe, the United States, China, and the Philippines. Approval for this research was obtained from the Institutional Review Boards of participating institutions (both US and foreign), and informed consent was obtained from parents of minor children and from all affected individuals old enough to give their own consent. Affected subjects were diagnosed as having cleft lip (CL) or cleft lip with cleft palate (CLP) based on physical exam. Individuals with other congenital anomalies, recognized syndromes, or developmental delays were excluded from this study.

### Regions selected and sequencing

This dataset was a targeted sequencing study covering 6.3Mb of the genome and was comprised of thirteen high-priority regions selected for sequencing in 2010 (Table S1), based on the best evidence from GWAS ^24, 36^, linkage studies ^23, 37, 38^, or candidate gene studies ^30^. As described previously, sequencing performed on Illumina multiplexed libraries constructed with 1 mg of native genomic DNA ^39^. Reads were mapped to the GRCh37 reference sequence by BWA v.0.5.912. *De novo* variant (DNV) calls were generated with Polymutt (v.0.11), which employed a likelihood-based framework that leveraged the parental genotype information to increase sensitivity and specificity when calling *de novo* variants. As part of the previous QC, identity by descent (IBD) was calculated, sites where 50% or more individuals had a false-positive filter flag were removed as were individual variant calls with a depth (DP) less than 7 or a genotype quality (GQ) less than 20.

### Functional annotation

DNVs were annotated to functional categories, such as predicted craniofacial enhancers and promoters, using published epigenetic marks and the derived chromatin states generated with ChromHMM in human embryonic craniofacial tissue ^35^. Additionally, DNVs were annotated to experimentally validated enhancers from previous research conducted in oral epithelium or palate mesenchyme cells ^40^, enhancers identified from genome-wide ChIP-seq experiments for the enhancer-associated p300 protein in mouse craniofacial tissue ^41^, and enhancers from publicly available databases like the VISTA Enhancer Browser (filtering for positive enhancers in the branchial arch, facial mesenchyme, and mesenchyme derived from neural crest)^42^. For any enhancer region with a DNV, we defined the topologically-associated domains (TADs) using data from cranial neural crest cells^35^. Gene expression for the genes within these DNV-containing TADs was then assessed with RNA-seq data generated from human cranial neural crest cells (GEO: GSM1817212, GSM1817213, GSM1817214, GSM1817215, GSM1817216, and GSM1817217) ^43, 44^. All variant calls and annotation regions were lifted over to the hg38 reference build for matching overlap of positions.

### Statistical analyses

#### De novo variants

For the hs1617 element that had more than one *de novo* variant, an excess burden of DNVs was calculated using Poisson distribution for the region size (N = 3,561 bp), the number of samples (N = 1,409), and the predicted mutation rate. The predicted mutation rate for this study was calculated based on the sequence of the enhancer and the summed trinucleotide mutation rate reported by Chen et al. ^45^. Because only one region was tested, p < 0.05 was considered statistically significant.

#### Rare variants

A burden of rare variants in the hs1617 region was tested using a rare variant extension of the transmission disequilibrium test (RV-TDT) ^46^ using published data from a whole-genome sequencing study of 759 trios with OFCs sequenced as part of the Gabriella Miller Kids First Pediatric Research Consortium^28, 47^. We considered this dataset complementary to the targeted sequencing study rather than an independent cohort due to some overlap between studies. Details of the sequencing and quality controls steps were previously described ^28, 47^. For this analysis, we retained only biallelic variants with a quality by depth (QD) ≥ 4 and a minor allele frequency (MAF) < 0.1% in any population as defined in gnomAD (v3.1.2). P < 0.05 was considered statistically significant.

#### Common variants

The association with common variants in hs1617 with OFCs was assessed with a previously published genome-wide meta-analysis that combined data from the Pittsburgh Orofacial Cleft (POFC) study and the GENEVA OFC study ^19^. The details of the original sample collection and genotype quality control (QC) have been described previously ^20, 36^, and the original studies recruited individuals with OFCs, their unaffected relatives, and unrelated controls (individuals with no known family history of OFCs or other craniofacial anomalies). From this data, three analysis groups were defined in the meta-analysis: a case-control group from the POFC data, an unrelated case-parent trio group from POFC data, and an unrelated case-parent trio group from GENEVA OFC data. There is likely some overlap in samples between the trios selected for targeted sequencing as the samples selected for SNP array. In all analysis groups, SNP markers showing deviation from Hardy-Weinberg equilibrium in unrelated European ancestry controls, MAF < 1%, or imputation INFO scores < 0.5 were filtered out of all subsequent analyses. In the case-control group, a logistic regression model was used to test for association with 18 PCs included as covariates to control for population stratification. In both groups of trios, a TDT was used to test for association. PLINK was used for all analyses ^48^. The results from the three analysis groups were combined in an inverse variance-weighted fixed-effects meta-analysis^49^, and SNPs with association p-values less than 5 X 10^−8^ were considered genome-wide significant. Regional association plots were visualized with LocusZoom ^50^.

#### Bioinformatic Analysis

For any predicted enhancer or promoter region that contained a DNV, transcription factor binding sites (TFBS) were predicted using TFBSTools ^51^ and the JASPAR database ^52^. The 50 base pairs surrounding the DNV were extracted from the reference sequence, and the analysis was conducted using both the reference sequence and the mutated sequence. We then selected predicted TFBS that both overlapped with the position of the DNV and differed between the reference and mutated sequence. TBFS were considered predicted if the p-value was significant after multiple test correction (p < 0.05/Number of TFBS that overlapped the DNV position). The TFBS were considered different between reference and mutated sequence if the TFBS was significant in one sequence and not the other.

### Functional studies of hs1617

Transgenic mouse embryos were generated as previously described^53^. Briefly, enhancer sequences (hs1617, hs1617.1 and hs1617.2) were synthesized by Twist Biosciences and cloned into the vector containing minimal Shh promoter, lacZ reporter gene and H11 locus homology arms (Addgene, 139098) using NEBuilder HiFi DNA Assembly Mix (NEB, E2621). The sequence of the resulting assembled plasmids was verified using long-read sequencing (Plasmidsaurus). All plasmids are available upon request. Fertilized embryos, from super-ovulated FVB females mated with FVB males, were collected from the oviducts. An injection mixture of Cas9 protein (Alt-R SpCas9 Nuclease V3, IDT, Cat#1081058, final concentration 20 ng/uL), hybridzed sgRNA against the H11 locus (Alt-R CRISPR-Cas9 tracrRNA, IDT, Cat#1072532 and Alt-R CRISPR-Cas9 locus targeting crRNA, gctgatggaacaggtaacaa, total final concentration 50 ng/μL) and the vector (12.5 ng/µL) was injected into the pronucleus of zygotes. Zygotes were cultured in M16 with amino acids at 37°C, 5% CO_2_ for 2 hours and then transferred into pseudopregnant CD-1 mice. Embryos were collected at E13.5 and stained for lacZ activity as previously described^53^. Briefly, embryos were dissected from the uterine horns (keeping the sacs attached) and washed in PBS. Embryos were fixed in 4% PFA for 45 mins for E13.5 embryos at room temperature then washed three times in embryo wash buffer (2 mM MgCl2, 0.02% NP-40 and 0.01% deoxycholate in PBS at pH 7.3). This was followed by overnight staining with X-gal (4 mM potassium ferricyanide, 4 mM potassium ferrocyanide, 1 mg/mL X-gal and 20 mM Tris pH 7.5 in embryo wash buffer) at room temperature. Stained embryos were washed three times in PBS and room temperature and embryos stored in 4% PFA at 4°C. Separately, embryo sacs were digested with DirectPCR Lysis Reagent (Viagen, 301-C) with Proteinase K added (final concentration 6 U/mL) to extract genomic DNA for genotyping. Genotyping PCR was performed to confirm integration at the H11 locus and assess for single and tandem construct insertions ^53^. Transgenic embryos were imaged using a Leica MZ16 microscope and Leica DFC420 digital camera and processed to remove background in Adobe Photoshop. Embryos for all regions tested are publicly available in the VISTA Enhancer Browser ^54, 55^ (https://enhancer.lbl.gov)

## Results

We reanalyzed *de novo* variants (DNVs) that were called in 1,409 trios as part of a targeted sequencing study ^30^ to assess if any were contained in newly predicted functional regions based on recent epigenetic or experimental data. The original study sequenced 13 regions and called 66 DNVs in known OFC-associated regions. When we reannotated the DNVs to their predicted functional regions (Table 1, Figure S1), most (N = 44) were in regions of low activity or in regions of repressed chromatin. However, the other 22 (33%) were in regions involved in transcription or regulation, including 16 that were in regions designated as enhancers and 1 that was in a bivalent promoter.

**Table 1:**
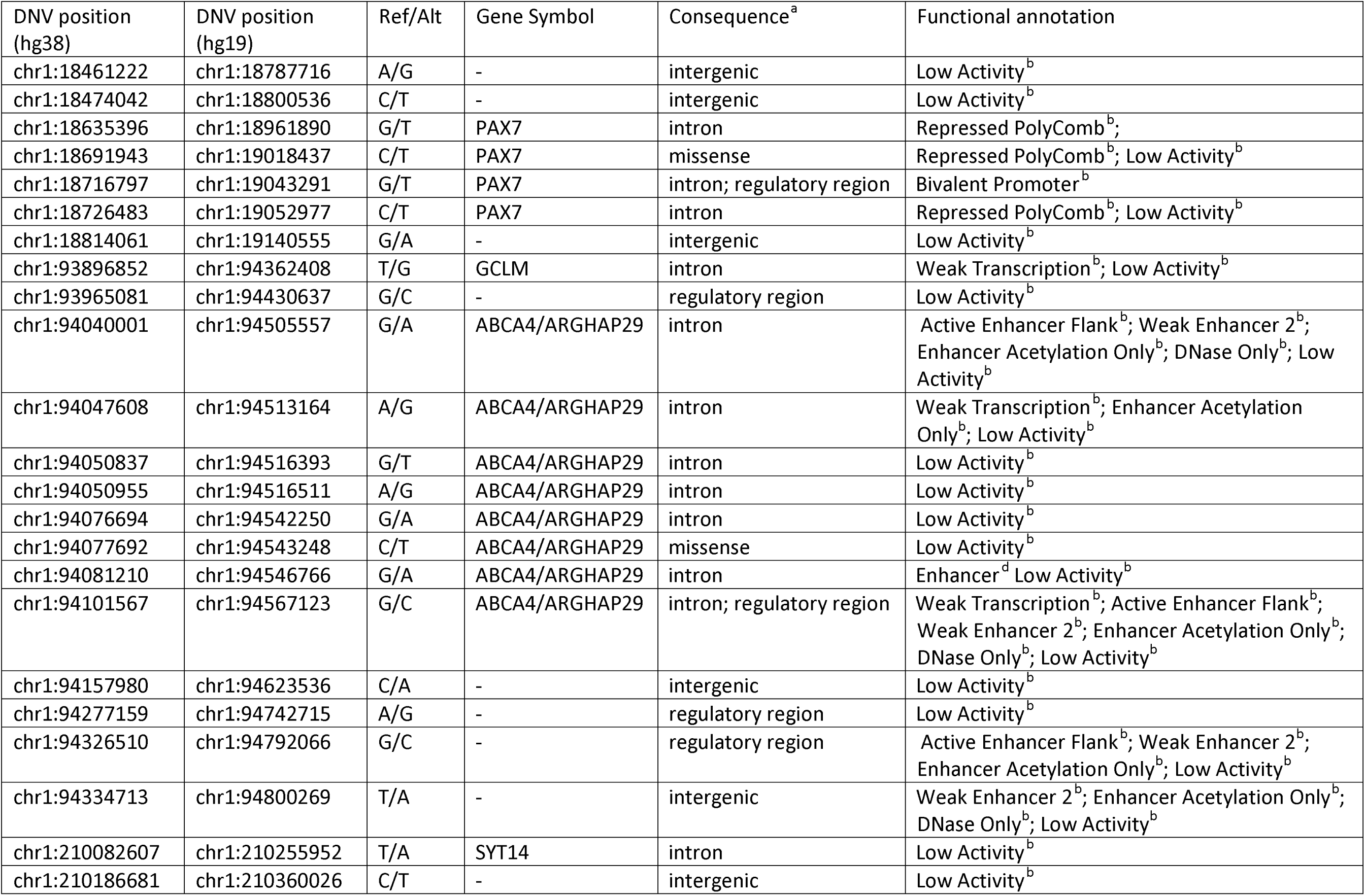
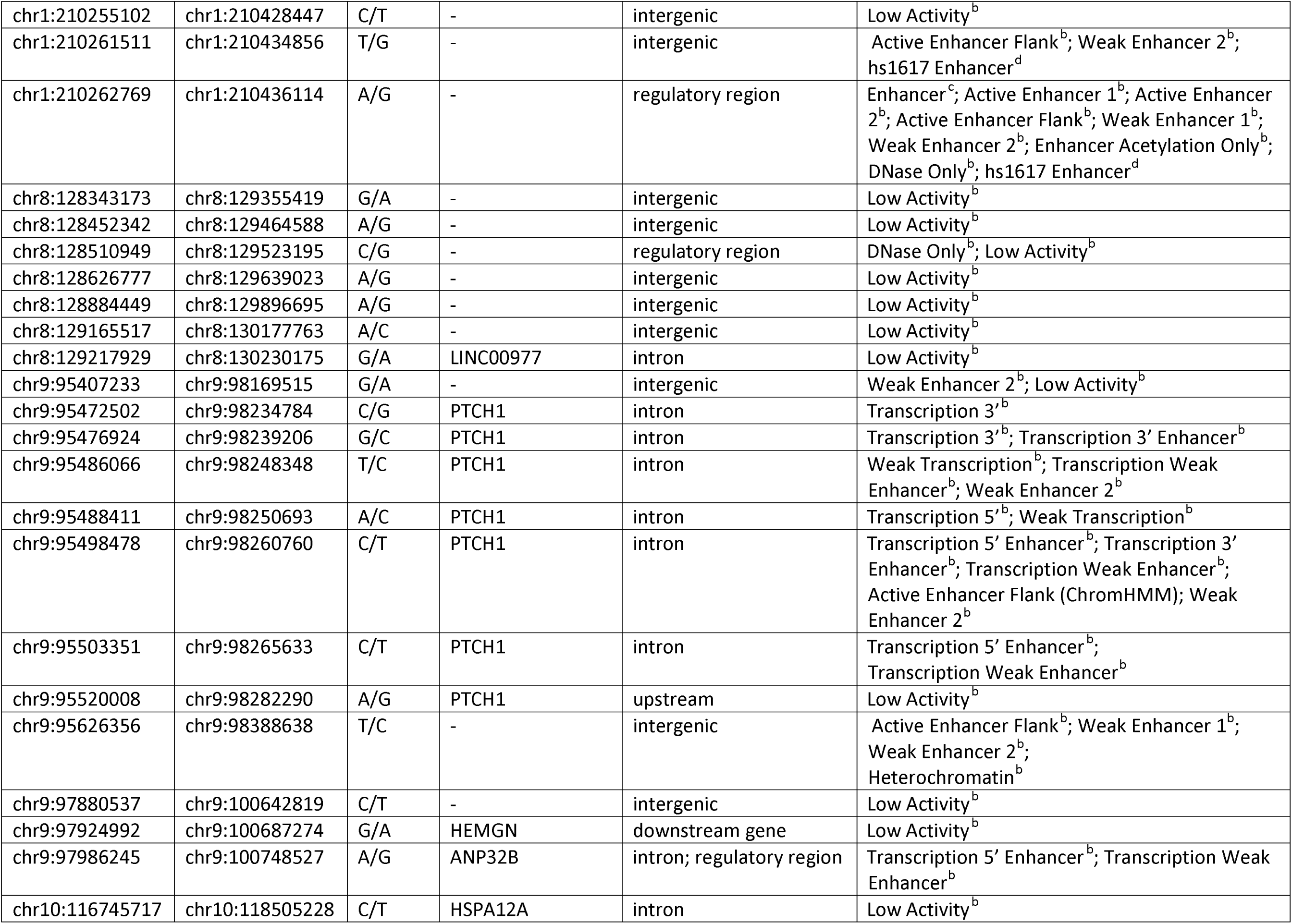
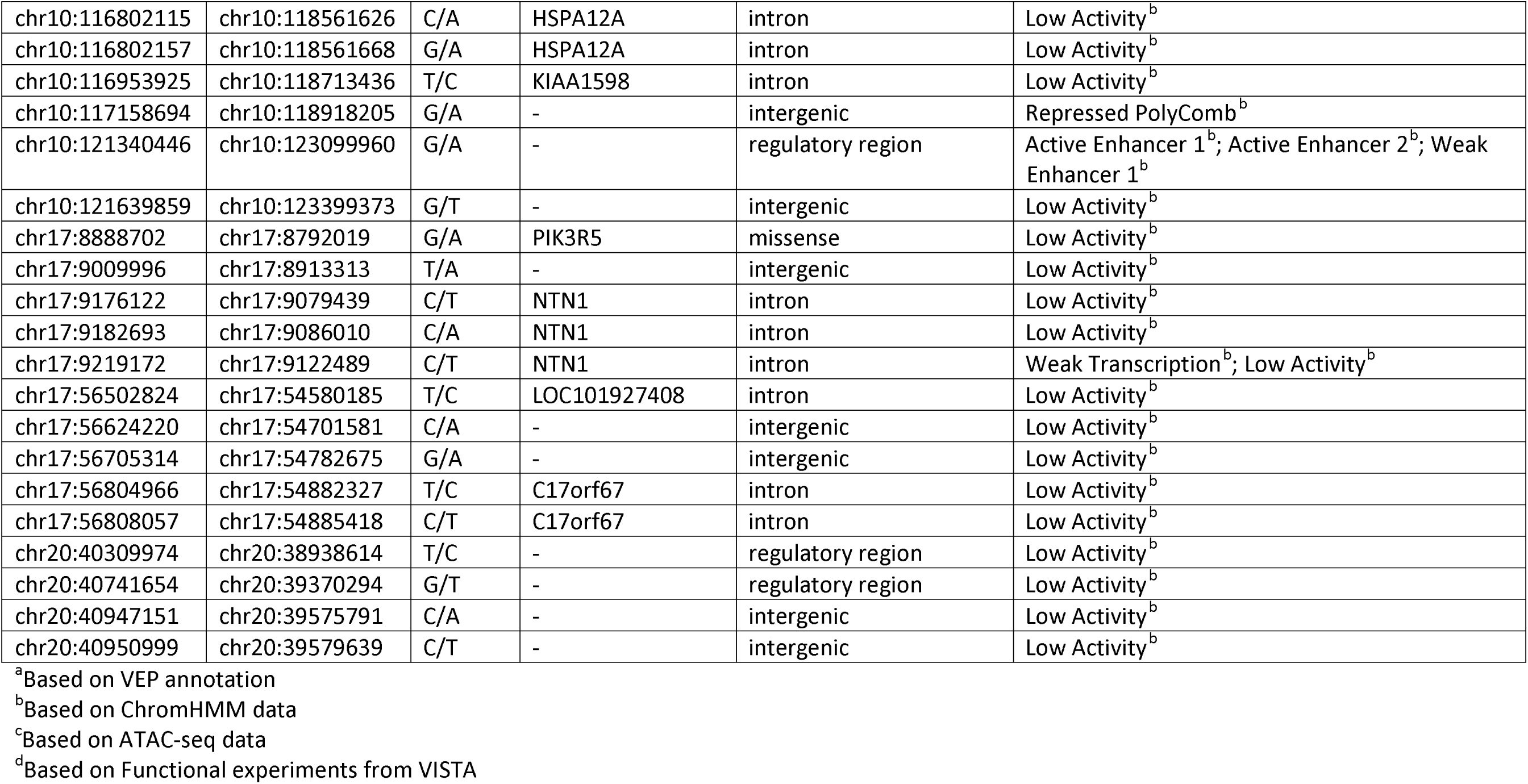
DNV positions and functional annotation.

| DNV position (hg38) | DNV position (hg19) | Ref/Alt | Gene Symbol | Consequence<sup>a</sup> | Functional annotation |
| --- | --- | --- | --- | --- | --- |
| chr1:18461222 | chr1:18787716 | A/G | - | intergenic | Low Activity<sup>b</sup> |
| chr1:18474042 | chr1:18800536 | C/T | - | intergenic | Low Activity<sup>b</sup> |
| chr1:18635396 | chr1:18961890 | G/T | PAX7 | intron | Repressed PolyComb<sup>b</sup>; |
| chr1:18691943 | chr1:19018437 | C/T | PAX7 | missense | Repressed PolyComb<sup>b</sup>; Low Activity<sup>b</sup> |
| chr1:18716797 | chr1:19043291 | G/T | PAX7 | intron; regulatory region | Bivalent Promoter<sup>b</sup> |
| chr1:18726483 | chr1:19052977 | C/T | PAX7 | intron | Repressed PolyComb<sup>b</sup>; Low Activity<sup>b</sup> |
| chr1:18814061 | chr1:19140555 | G/A | - | intergenic | Low Activity<sup>b</sup> |
| chr1:93896852 | chr1:94362408 | T/G | GCLM | intron | Weak Transcription<sup>b</sup>; Low Activity<sup>b</sup> |
| chr1:93965081 | chr1:94430637 | G/C | - | regulatory region | Low Activity<sup>b</sup> |
| chr1:94040001 | chr1:94505557 | G/A | ABCA4/ARGHAP29 | intron | Active Enhancer Flank<sup>b</sup>; Weak Enhancer 2<sup>b</sup>; Enhancer Acetylation Only<sup>b</sup>; DNase Only<sup>b</sup>; Low Activity<sup>b</sup> |
| chr1:94047608 | chr1:94513164 | A/G | ABCA4/ARGHAP29 | intron | Weak Transcription<sup>b</sup>; Enhancer Acetylation Only<sup>b</sup>; Low Activity<sup>b</sup> |
| chr1:94050837 | chr1:94516393 | G/T | ABCA4/ARGHAP29 | intron | Low Activity<sup>b</sup> |
| chr1:94050955 | chr1:94516511 | A/G | ABCA4/ARGHAP29 | intron | Low Activity<sup>b</sup> |
| chr1:94076694 | chr1:94542250 | G/A | ABCA4/ARGHAP29 | intron | Low Activity<sup>b</sup> |
| chr1:94077692 | chr1:94543248 | C/T | ABCA4/ARGHAP29 | missense | Low Activity<sup>b</sup> |
| chr1:94081210 | chr1:94546766 | G/A | ABCA4/ARGHAP29 | intron | Enhancer<sup>d</sup> Low Activity<sup>b</sup> |
| chr1:94101567 | chr1:94567123 | G/C | ABCA4/ARGHAP29 | intron; regulatory region | Weak Transcription<sup>b</sup>; Active Enhancer Flank<sup>b</sup>; Weak Enhancer 2<sup>b</sup>; Enhancer Acetylation Only<sup>b</sup>; DNase Only<sup>b</sup>; Low Activity<sup>b</sup> |
| chr1:94157980 | chr1:94623536 | C/A | - | intergenic | Low Activity<sup>b</sup> |
| chr1:94277159 | chr1:94742715 | A/G | - | regulatory region | Low Activity<sup>b</sup> |
| chr1:94326510 | chr1:94792066 | G/C | - | regulatory region | Active Enhancer Flank<sup>b</sup>; Weak Enhancer 2<sup>b</sup>; Enhancer Acetylation Only<sup>b</sup>; Low Activity<sup>b</sup> |
| chr1:94334713 | chr1:94800269 | T/A | - | intergenic | Weak Enhancer 2<sup>b</sup>; Enhancer Acetylation Only<sup>b</sup>; DNase Only<sup>b</sup>; Low Activity<sup>b</sup> |
| chr1:210082607 | chr1:210255952 | T/A | SYT14 | intron | Low Activity<sup>b</sup> |
| chr1:210186681 | chr1:210360026 | C/T | - | intergenic | Low Activity<sup>b</sup> |
| chr1:210255102 | chr1:210428447 | C/T | - | intergenic | Low Activity<sup>b</sup> |
| chr1:210261511 | chr1:210434856 | T/G | - | intergenic | Active Enhancer Flank<sup>b</sup>; Weak Enhancer 2<sup>b</sup>; hs1617 Enhancer<sup>d</sup> |
| chr1:210262769 | chr1:210436114 | A/G | - | regulatory region | Enhancer<sup>c</sup>; Active Enhancer 1<sup>b</sup>; Active Enhancer 2<sup>b</sup>; Active Enhancer Flank<sup>b</sup>; Weak Enhancer 1<sup>b</sup>; Weak Enhancer 2<sup>b</sup>; Enhancer Acetylation Only<sup>b</sup>; DNase Only<sup>b</sup>; hs1617 Enhancer<sup>d</sup> |
| chr8:128343173 | chr8:129355419 | G/A | - | intergenic | Low Activity<sup>b</sup> |
| chr8:128452342 | chr8:129464588 | A/G | - | intergenic | Low Activity<sup>b</sup> |
| chr8:128510949 | chr8:129523195 | C/G | - | regulatory region | DNase Only<sup>b</sup>; Low Activity<sup>b</sup> |
| chr8:128626777 | chr8:129639023 | A/G | - | intergenic | Low Activity<sup>b</sup> |
| chr8:128884449 | chr8:129896695 | A/G | - | intergenic | Low Activity<sup>b</sup> |
| chr8:129165517 | chr8:130177763 | A/C | - | intergenic | Low Activity<sup>b</sup> |
| chr8:129217929 | chr8:130230175 | G/A | LINC00977 | intron | Low Activity<sup>b</sup> |
| chr9:95407233 | chr9:98169515 | G/A | - | intergenic | Weak Enhancer 2<sup>b</sup>; Low Activity<sup>b</sup> |
| chr9:95472502 | chr9:98234784 | C/G | PTCH1 | intron | Transcription 3'<sup>b</sup> |
| chr9:95476924 | chr9:98239206 | G/C | PTCH1 | intron | Transcription 3'<sup>b</sup>; Transcription 3' Enhancer<sup>b</sup> |
| chr9:95486066 | chr9:98248348 | T/C | PTCH1 | intron | Weak Transcription<sup>b</sup>; Transcription Weak Enhancer<sup>b</sup>; Weak Enhancer 2<sup>b</sup> |
| chr9:95488411 | chr9:98250693 | A/C | PTCH1 | intron | Transcription 5'<sup>b</sup>; Weak Transcription<sup>b</sup> |
| chr9:95498478 | chr9:98260760 | C/T | PTCH1 | intron | Transcription 5' Enhancer<sup>b</sup>; Transcription 3' Enhancer<sup>b</sup>; Transcription Weak Enhancer<sup>b</sup>; Active Enhancer Flank (ChromHMM); Weak Enhancer 2<sup>b</sup> |
| chr9:95503351 | chr9:98265633 | C/T | PTCH1 | intron | Transcription 5' Enhancer<sup>b</sup>; Transcription Weak Enhancer<sup>b</sup> |
| chr9:95520008 | chr9:98282290 | A/G | PTCH1 | upstream | Low Activity<sup>b</sup> |
| chr9:95626356 | chr9:98388638 | T/C | - | intergenic | Active Enhancer Flank<sup>b</sup>; Weak Enhancer 1<sup>b</sup>; Weak Enhancer 2<sup>b</sup>; Heterochromatin<sup>b</sup> |
| chr9:97880537 | chr9:100642819 | C/T | - | intergenic | Low Activity<sup>b</sup> |
| chr9:97924992 | chr9:100687274 | G/A | HEMGN | downstream gene | Low Activity<sup>b</sup> |
| chr9:97986245 | chr9:100748527 | A/G | ANP32B | intron; regulatory region | Transcription 5' Enhancer<sup>b</sup>; Transcription Weak Enhancer<sup>b</sup> |
| chr10:116745717 | chr10:118505228 | C/T | HSPA12A | intron | Low Activity<sup>b</sup> |
| chr10:116802115 | chr10:118561626 | C/A | HSPA12A | intron | Low Activity<sup>b</sup> |
| chr10:116802157 | chr10:118561668 | G/A | HSPA12A | intron | Low Activity<sup>b</sup> |
| chr10:116953925 | chr10:118713436 | T/C | KIAA1598 | intron | Low Activity<sup>b</sup> |
| chr10:117158694 | chr10:118918205 | G/A | - | intergenic | Repressed PolyComb<sup>b</sup> |
| chr10:121340446 | chr10:123099960 | G/A | - | regulatory region | Active Enhancer 1<sup>b</sup>; Active Enhancer 2<sup>b</sup>; Weak Enhancer 1<sup>b</sup> |
| chr10:121639859 | chr10:123399373 | G/T | - | intergenic | Low Activity<sup>b</sup> |
| chr17:8888702 | chr17:8792019 | G/A | PIK3R5 | missense | Low Activity<sup>b</sup> |
| chr17:9009996 | chr17:8913313 | T/A | - | intergenic | Low Activity<sup>b</sup> |
| chr17:9176122 | chr17:9079439 | C/T | NTN1 | intron | Low Activity<sup>b</sup> |
| chr17:9182693 | chr17:9086010 | C/A | NTN1 | intron | Low Activity<sup>b</sup> |
| chr17:9219172 | chr17:9122489 | C/T | NTN1 | intron | Weak Transcription<sup>b</sup>; Low Activity<sup>b</sup> |
| chr17:56502824 | chr17:54580185 | T/C | LOC101927408 | intron | Low Activity<sup>b</sup> |
| chr17:56624220 | chr17:54701581 | C/A | - | intergenic | Low Activity<sup>b</sup> |
| chr17:56705314 | chr17:54782675 | G/A | - | intergenic | Low Activity<sup>b</sup> |
| chr17:56804966 | chr17:54882327 | T/C | C17orf67 | intron | Low Activity<sup>b</sup> |
| chr17:56808057 | chr17:54885418 | C/T | C17orf67 | intron | Low Activity<sup>b</sup> |
| chr20:40309974 | chr20:38938614 | T/C | - | regulatory region | Low Activity<sup>b</sup> |
| chr20:40741654 | chr20:39370294 | G/T | - | regulatory region | Low Activity<sup>b</sup> |
| chr20:40947151 | chr20:39575791 | C/A | - | intergenic | Low Activity<sup>b</sup> |
| chr20:40950999 | chr20:39579639 | C/T | - | intergenic | Low Activity<sup>b</sup> |

<sup>a</sup>Based on VEP annotation
<sup>b</sup>Based on ChromHMM data
<sup>c</sup>Based on ATAC-seq data
<sup>d</sup>Based on Functional experiments from VISTA

To assess whether any functional region had a burden of DNVs compared to what was expected by chance, we tested for enrichment of DNVs. We found one enhancer, hs1617, had 2 DNVs within it (chr1:210261511:T:G and chr1:210262769:A:G), which was significantly more than expected by chance given the size and sequence context of the enhancer (p = 0.0017). This enhancer was within a large (∼400kb) region that was associated with OFCs (lead SNP p-value = 2.78 × 10^−13^) in a previous meta-analysis ^19^, particularly with CLP (Figure S2) and is in a large LD block with *IRF6,* a well-known clefting gene ^17, 20, 56–61^. We did not see a burden of rare, inherited genetic variants in this enhancer associated with OFCs in an RV-TDT with 20 variants (not containing the 2 DNVs) identified in a separate cohort with genome sequencing data (p = 0.39).

Because DNVs in enhancer regions could impact transcription factor binding sites (TFBSs) and thus impact gene regulation, we then further investigated all DNVs that were within enhancers. For the 2 DNVs in hs1617, the variants are expected to disrupt the predicted binding sites for STAT1 and STAT3 and create binding sites for PAX6, ZBTB7A, and MZF1 (Table 2, Table S3-S4). The topologically-associated domain (TAD) around hs1617 contains *SERTAD4*, *HHAT*, and *IRF6* (Figure 1). Both *SERTAD4* and *HHAT* were in the top 20^th^ percentile of genes expressed in neural crest cell lines ^62^. To follow up on our *in silico* predictions for the possible effects of these two hs1617 DNVs, we tested whether this enhancer had craniofacial activity in a transgenic reporter mouse model. Enhancers displayed chondrogenic activity at embryonic day 13 as well as activity in the palatal shelves (Figure S3). However, the presence of either DNV did not lead to a detectable alteration in enhancer activity (Figure S3). We note that the other DNVs located within enhancers were also in TADs containing genes in the top 20^th^ percentile of expression in developing human craniofacial tissue^62^ (Figure S4) like *ARHGAP29*, *PTCH1*, and *FGFR2*, and the DNVs within these enhancers were also predicted to lead to altered TFBSs (Table S5-S19), indicating that these DNVs could be functionally relevant even if no single element reached statistical significance in this cohort.

**Figure 1:**
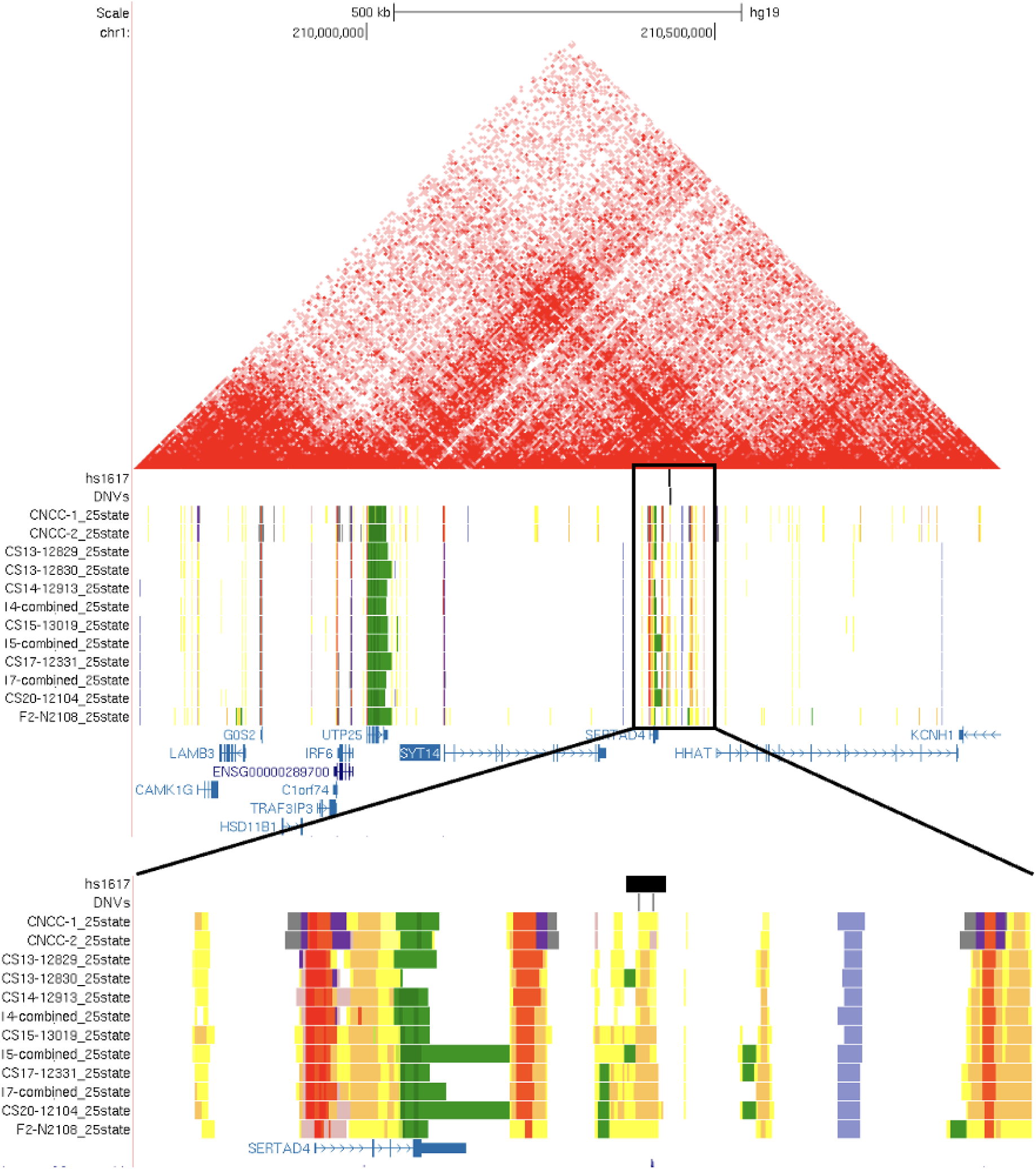
Multiple DNVs in hs1617. UCSC Genome Browser (hg38) view^35^ of region around the hs1617 element including the predicted TAD in cranial neural crest cells. Two of the DNVs in this study were within the hs1617 element.

**Table 2:**
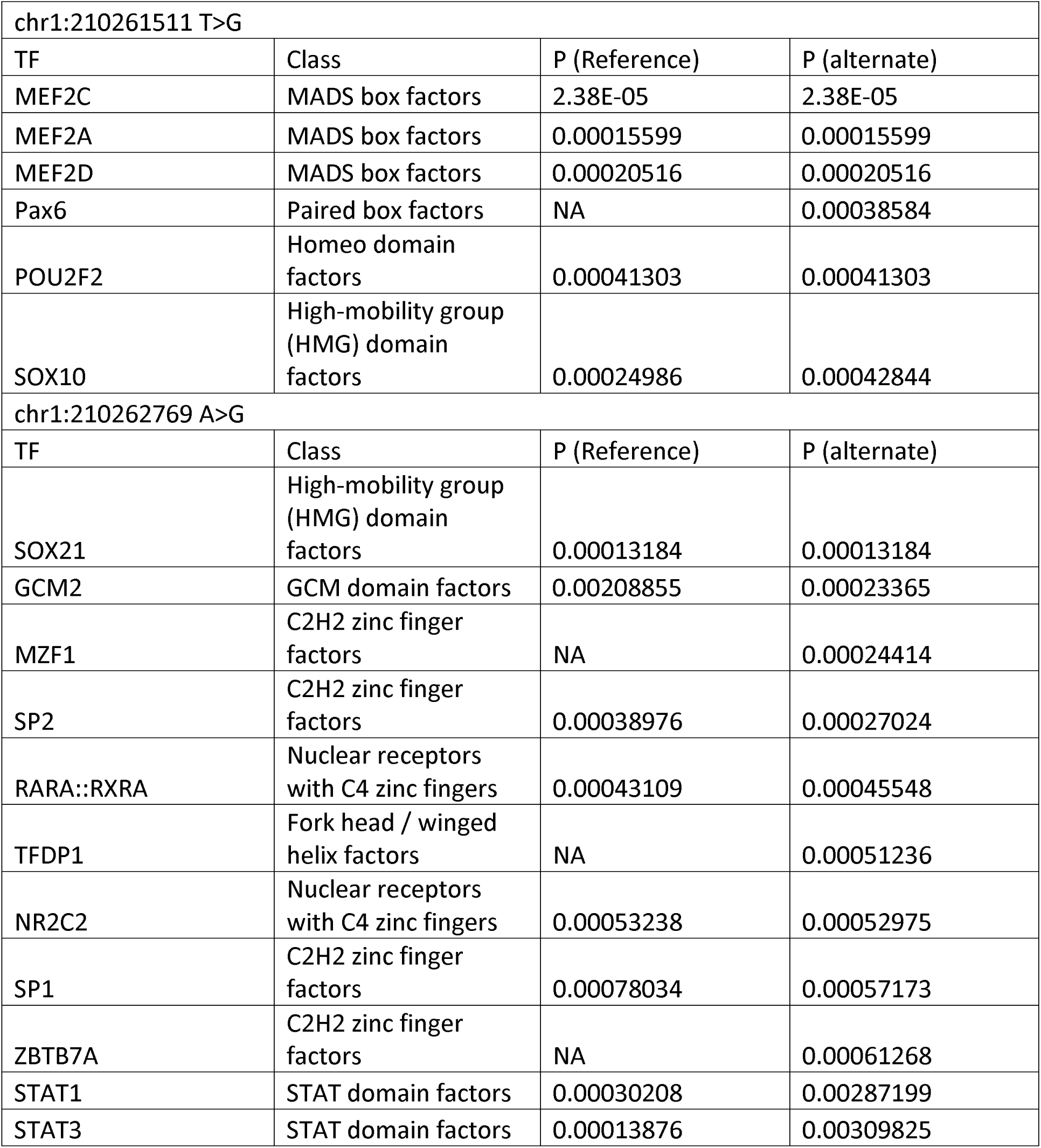
Statistically significant JASPAR predictions for DNVs in hs1617.

| chr1:210261511 T>G |  |  |  |
| --- | --- | --- | --- |
| TF | Class | P (Reference) | P (alternate) |
| MEF2C | MADS box factors | 2.38E-05 | 2.38E-05 |
| MEF2A | MADS box factors | 0.00015599 | 0.00015599 |
| MEF2D | MADS box factors | 0.00020516 | 0.00020516 |
| Pax6 | Paired box factors | NA | 0.00038584 |
| POU2F2 | Homeo domain factors | 0.00041303 | 0.00041303 |
| SOX10 | High-mobility group (HMG) domain factors | 0.00024986 | 0.00042844 |
| chr1:210262769 A>G |  |  |  |
| TF | Class | P (Reference) | P (alternate) |
| SOX21 | High-mobility group (HMG) domain factors | 0.00013184 | 0.00013184 |
| GCM2 | GCM domain factors | 0.00208855 | 0.00023365 |
| MZF1 | C2H2 zinc finger factors | NA | 0.00024414 |
| SP2 | C2H2 zinc finger factors | 0.00038976 | 0.00027024 |
| RARA::RXRA | Nuclear receptors with C4 zinc fingers | 0.00043109 | 0.00045548 |
| TFDP1 | Fork head / winged helix factors | NA | 0.00051236 |
| NR2C2 | Nuclear receptors with C4 zinc fingers | 0.00053238 | 0.00052975 |
| SP1 | C2H2 zinc finger factors | 0.00078034 | 0.00057173 |
| ZBTB7A | C2H2 zinc finger factors | NA | 0.00061268 |
| STAT1 | STAT domain factors | 0.00030208 | 0.00287199 |
| STAT3 | STAT domain factors | 0.00013876 | 0.00309825 |

## Discussion

While there have been previous studies of both coding *de novo variant*s (DNVs) ^28^ and a DNV in a regulatory region known at the time ^30^, the effect of DNVs in non-coding, regulatory regions have been less studied. One of the primary limitations for studying this in OFCs has been the identification of regulatory regions relevant in developing craniofacial tissue. However, more recent advances have allowed the use of epigenetics to identify potential regulatory regions for craniofacial development ^35^. In this study, we re-analyzed an existing data set of DNVs from a targeting sequencing study of 1,409 trios to assess whether any DNVs fell within predicted regulatory regions. We found 2 DNVs in the hs1617 enhancer region, which was significantly more than expected based on a sequence-conscious background mutation rate. This was the only element with more than 1 DNV within it, and both variants were predicted to alter transcription factor binding sites (TFBSs) for Pax6, ZBTB7A, STAT1, and STAT3. Pax6 has been associated with OFCs ^63^, variants in ZBTB7A have caused facial dysmorphism and macrocephaly ^64^, and STAT3 is involved in osteogenesis ^65^. This is consistent with the hypothesis that deleterious variation in regulatory regions can cause disease by impacting how TFs bind and regulate gene expression.

The hs1617 element is closest to *SERTAD4*, with *IRF6* and *HHAT* also being within the topologically-associated domain (TAD). While *IRF6* had the lowest expression of three genes, it has a well-documented association with both nonsyndromic OFCs ^17, 20, 26, 36, 66, 67^, and causes Van der Woude Syndrome (MIM# 119300), the most common form of syndromic OFCs that presents with an OFC and lip pits ^57, 59–61, 68, 69^. While their association with OFCs is not as well studied, *SERTAD4* and *HHAT* had higher expression in neural crest cell lines. While not much is known about the gene’s function, *SERTAD4* has been associated with nonsyndromic OFCs previously ^70, 71^, as has *HHAT* ^72, 73^. The is also evidence that homozygous missense variants in *HHAT* cause Nivelon-Nivelon-Mabille syndrome (which involves microcephaly and skeletal dysplasia) and other craniofacial defects ^74, 75^. Therefore, the genes regulated by the hs1617 element are relevant to craniofacial development, although more mechanistic work would be needed to directly test the effect that these variants have on gene expression and to determine which gene or genes hs1617 regulates.

By integrating newly predicted regulatory regions, this study builds on the previous work done in this dataset. In the original study, 3 of the DNVs were reported to be missense variants and 11 were reported to be in regulatory regions identified by ENCODE, with one of the regulatory variants near *FGFR2* being functional based on their follow-up experimental data ^30^. In this study, beyond finding the burden of DNVs in hs1617, which was not previously reported, we also found that one third of the DNVs identified were in either transcribed regions or regulatory regions, with 16 being in predicted enhancers and 1 being in a predicted bivalent promoter. These are potential candidates for functional studies to assess their effect on OFCs that were missed in the original analysis because of the limited data on functional regions for craniofacial tissue. This also demonstrates the value of secondary analyses with publicly available datasets.

This study is limited by the use of targeted sequencing. At the time of the original study, genome sequencing was in its infancy and not feasible at the scale necessary for studying complex traits. Reanalysis is therefore limited to original targeted design and does not allow for us to identify any novel regions associated with OFCs. There are certainly other regulatory elements in the genome that are associated with OFCs and could have a similar burden of DNVs, which will need to be identified through the growing datasets of genome sequencing for OFCs. In addition, further studies on regulatory elements and target gene expression genome-wide would help expand the interpretation of non-coding DNVs.

In conclusion, we found evidence for a burden of DNVs in the hs1617 enhancer in OFC cases. These DNVs were predicted to alter TFBSs near genes that are highly expressed in neural crest cells and associated with OFCs, providing further evidence that variation in regulatory regions is important for the genetic architecture of OFCs.

## Acknowledgments

The authors thank the collaborators, staff, and participating families for their important contributions to this study. This work was supported by grants from the National Institutes of Health (NIH) including: F32-DE032260 [SWC], R01-DE027362 [EJL], R03-DE030118 [EJL], U01-HG005925[JCM], R37-DE08559[JCM].

## Data Availability

The dbGaP accession number for the sequences reported in this paper is phs000625.v1.p1.

## Supplementary Tables

**Table S1:** Number of trios by ancestry and cleft type in targeted sequencing study.

|  | Number of trios with Asian ancestry | Number of trios with European ancestry | Total number of trios |
| --- | --- | --- | --- |
| Cleft lip | 286 | 102 | 388 |
| Cleft lip and palate | 729 | 257 | 986 |
| Cleft lip and cleft lip and palate | 19 | 16 | 35 |
| Total | 1034 | 375 | 1409 |

**Table S2:**
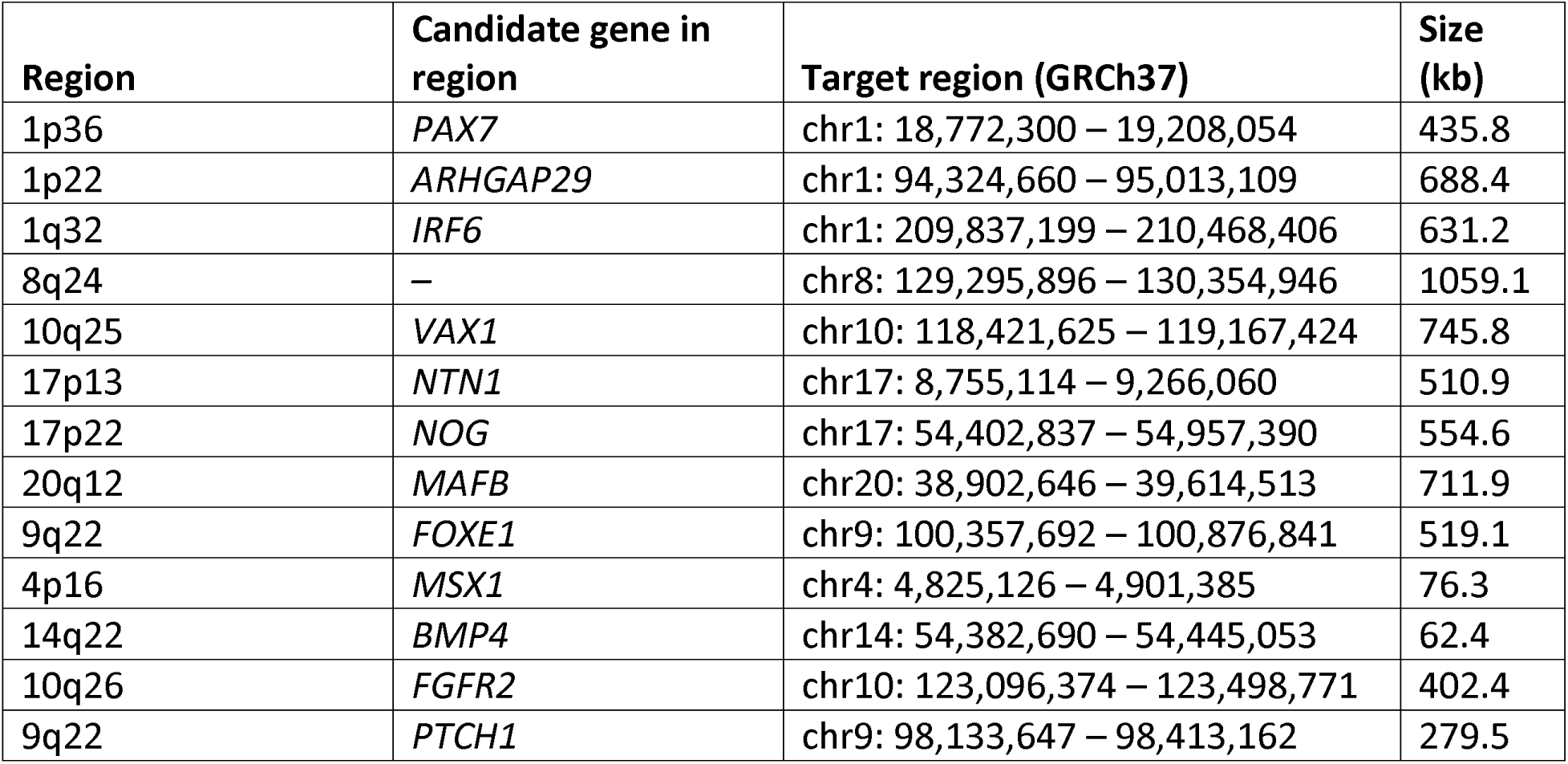
Regions selected for sequencing.

| Region | Candidate gene in region | Target region (GRCh37) | Size (kb) |
| --- | --- | --- | --- |
| 1p36 | *PAX7* | chr1: 18,772,300 – 19,208,054 | 435.8 |
| 1p22 | *ARHGAP29* | chr1: 94,324,660 – 95,013,109 | 688.4 |
| 1q32 | *IRF6* | chr1: 209,837,199 – 210,468,406 | 631.2 |
| 8q24 | – | chr8: 129,295,896 – 130,354,946 | 1059.1 |
| 10q25 | *VAX1* | chr10: 118,421,625 – 119,167,424 | 745.8 |
| 17p13 | *NTN1* | chr17: 8,755,114 – 9,266,060 | 510.9 |
| 17p22 | *NOG* | chr17: 54,402,837 – 54,957,390 | 554.6 |
| 20q12 | *MAFB* | chr20: 38,902,646 – 39,614,513 | 711.9 |
| 9q22 | *FOXE1* | chr9: 100,357,692 – 100,876,841 | 519.1 |
| 4p16 | *MSX1* | chr4: 4,825,126 – 4,901,385 | 76.3 |
| 14q22 | *BMP4* | chr14: 54,382,690 – 54,445,053 | 62.4 |
| 10q26 | *FGFR2* | chr10: 123,096,374 – 123,498,771 | 402.4 |
| 9q22 | *PTCH1* | chr9: 98,133,647 – 98,413,162 | 279.5 |

Table S3: JASPAR results for chr1:210261511 T>G

Table S4: JASPAR results for chr1:210262769 A>G

Table S5-S19: JASPAR results for enhancer and promoter regions

## Supplementary Figures

**Figure S1:**
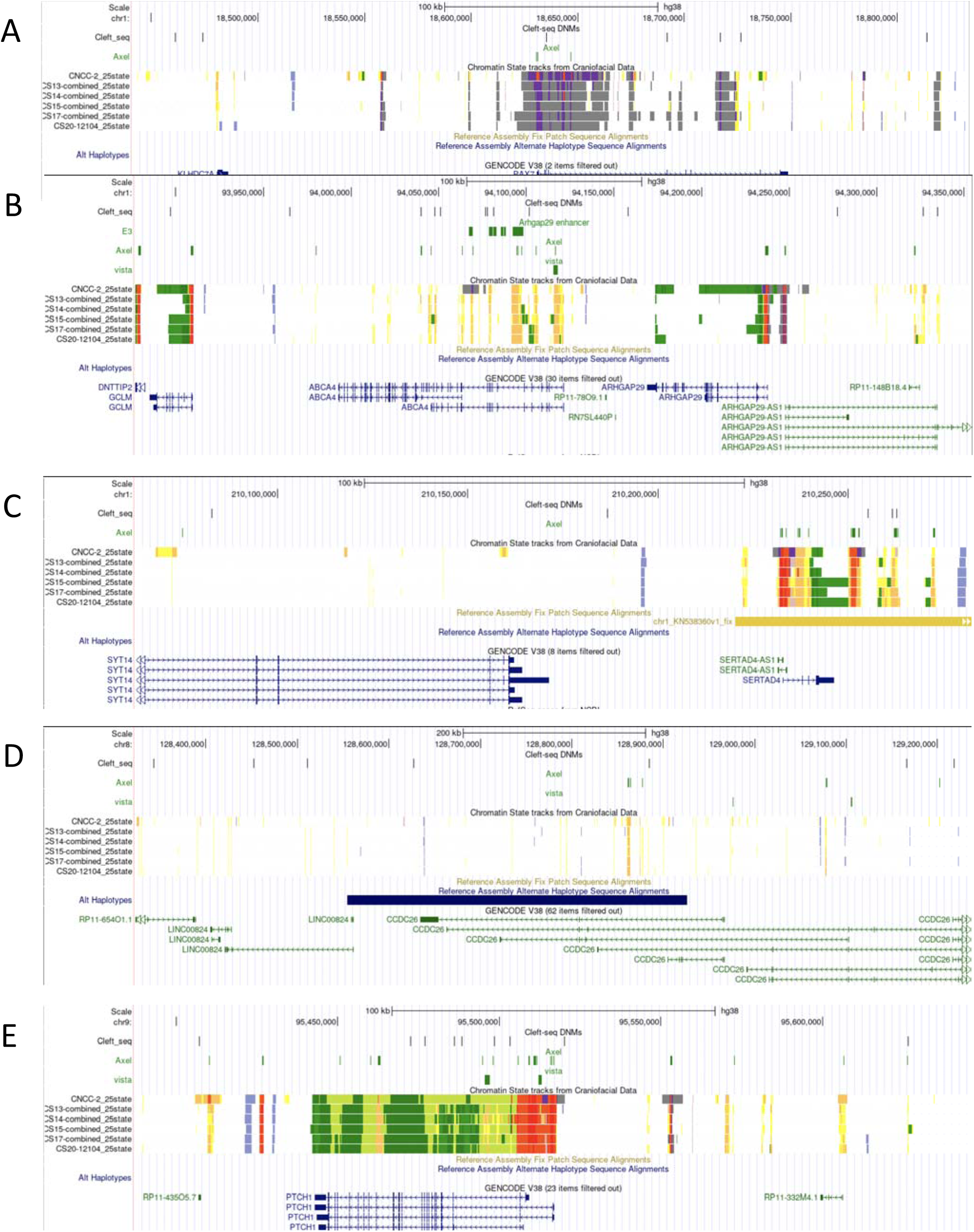
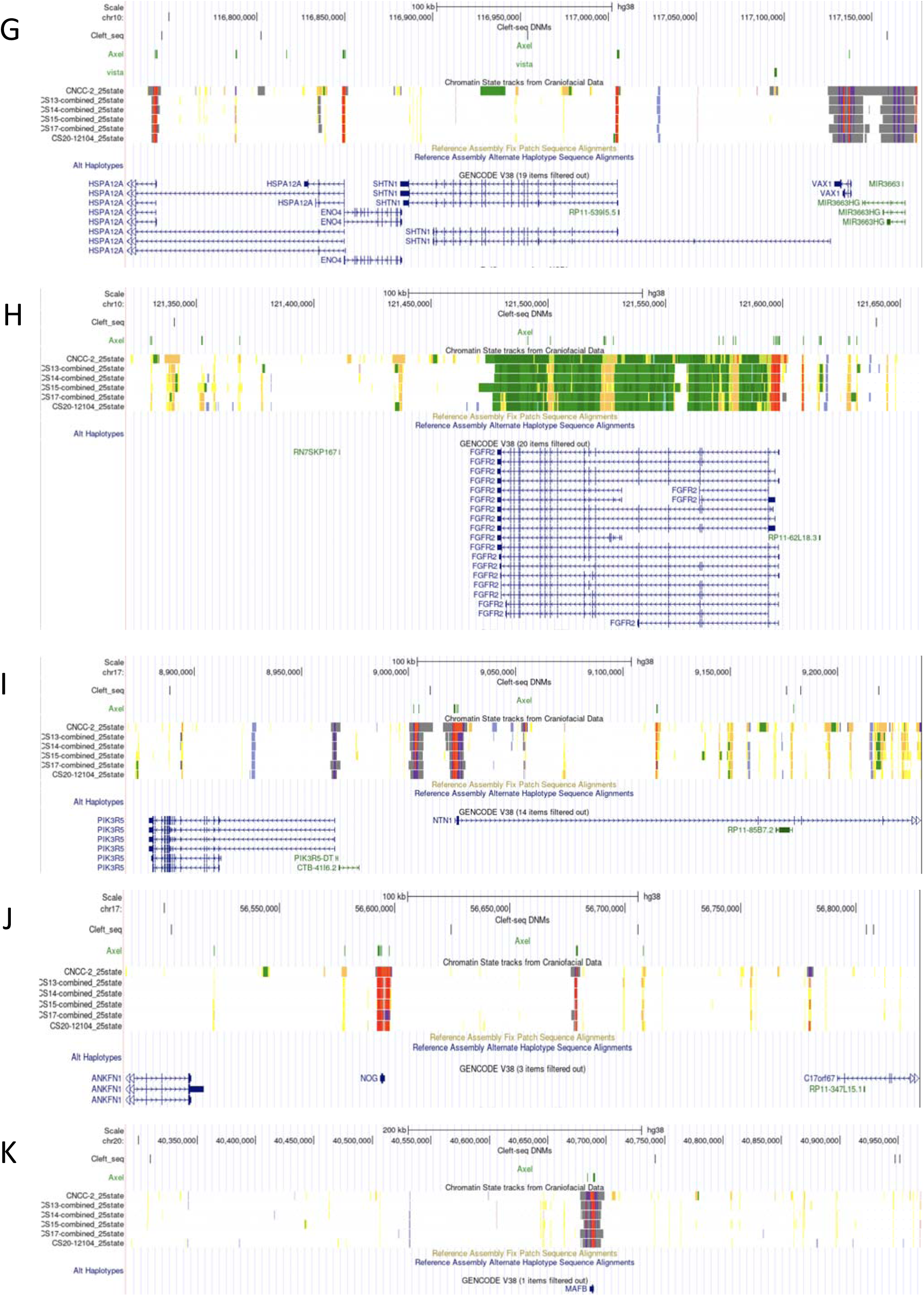
Regions for all DNVs. UCSC Genome Browser (hg38) view of PAX7 (**A**), ARHGAP29 (**B**), IRF6 (**C**), 8q24 (**D**), PTCH1 (**E**), FOXE1 (**F**), VAX1 (**G**), FGFR2 (**H**), NTN1 (**I**), NOG **(J**), MAFB (**K**) showing the position of the DNV found in this study with predicted functional regions.

**Figure S2:**
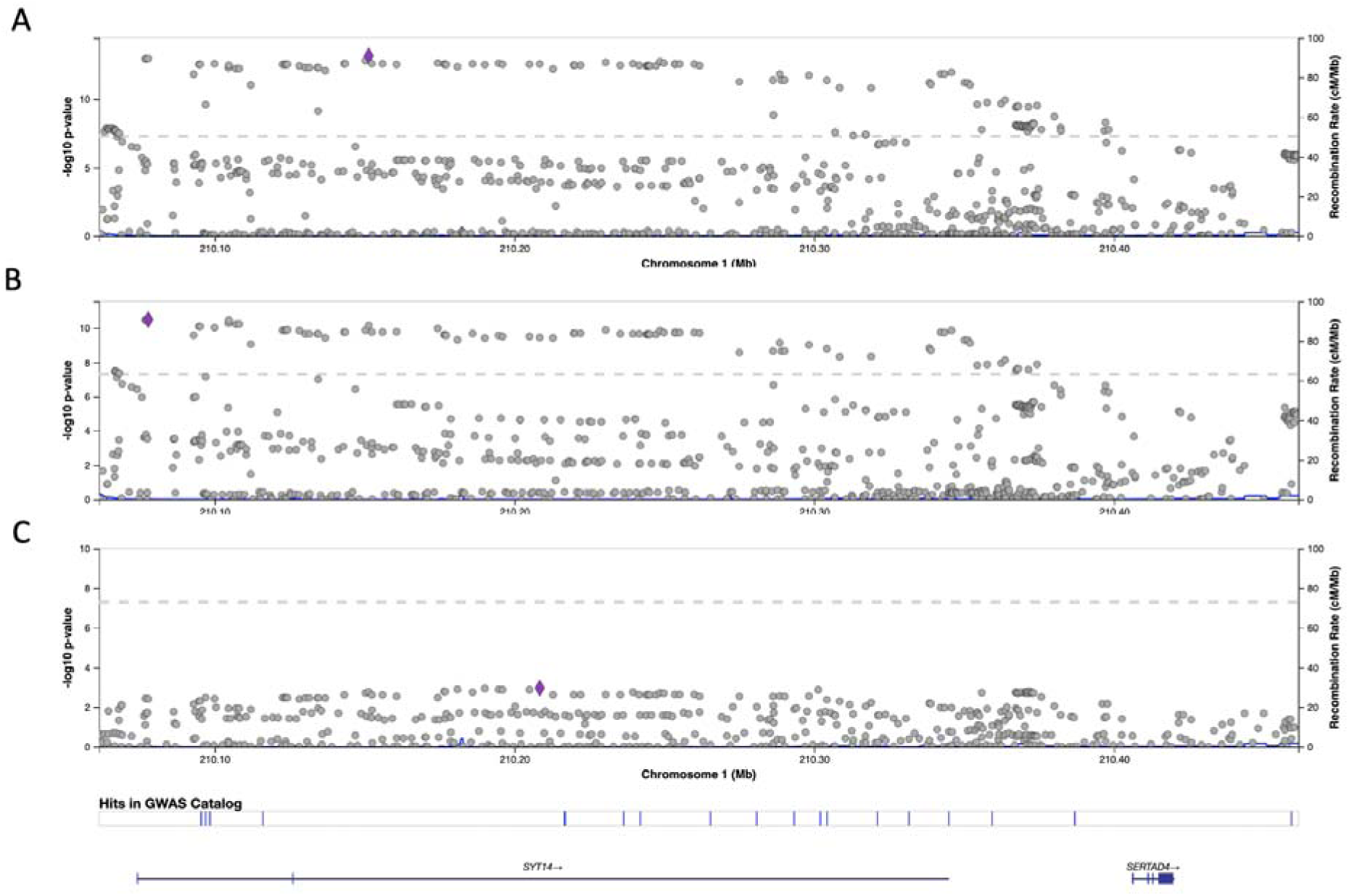
LocusZoom plot of hs1617 region. LocusZoom plots ^50^ for the hs1617 region showing the association of common variants in cleft lip with or without cleft palate (**A**), cleft lip and palate only (**B**), and cleft lip only (**C**) from a multi-ancestry meta-analysis.

**Figure S3:**
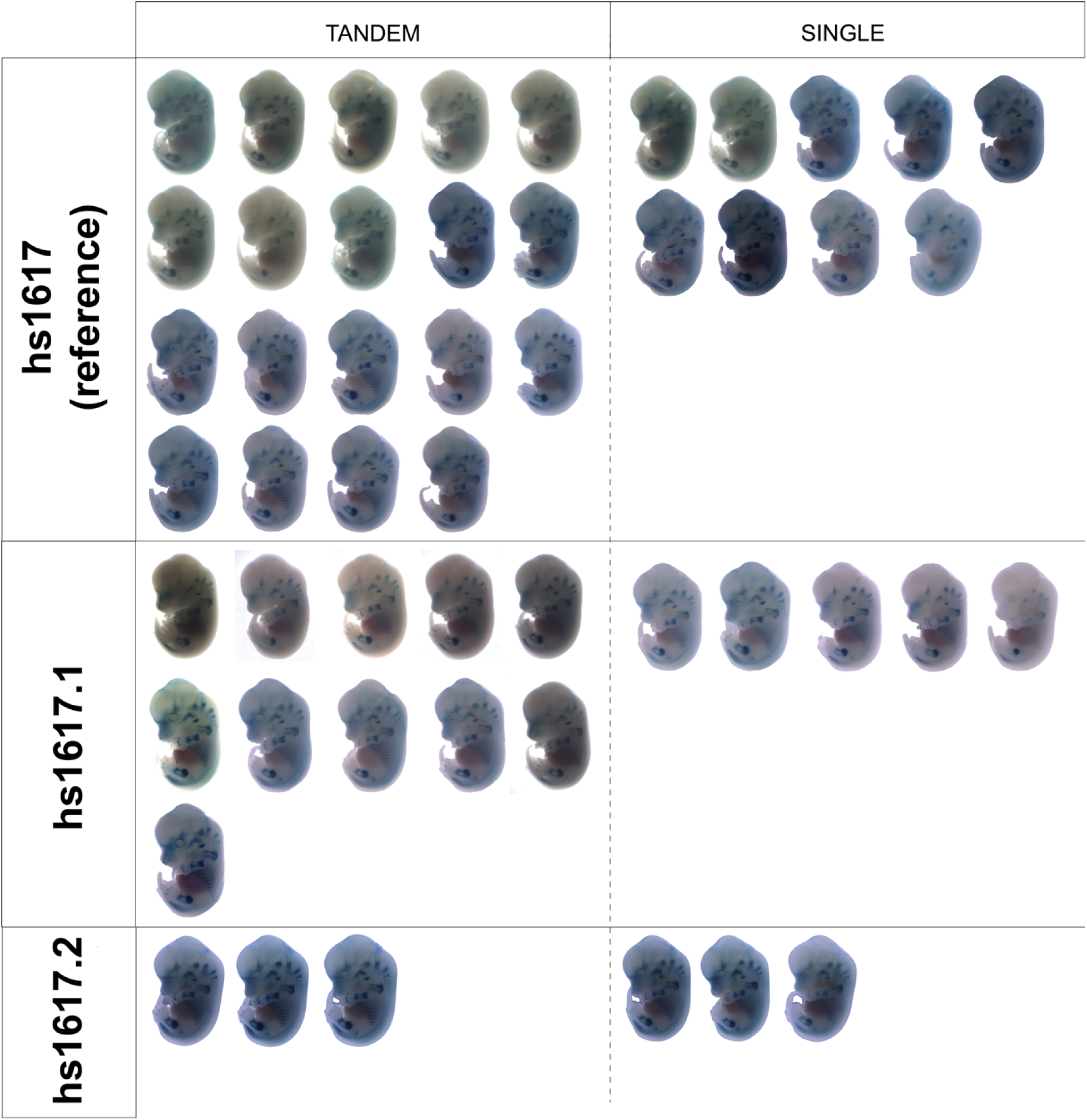
Functional testing of hs1617. Whole-mount transgenic reporter mice at embryonic day 13.5 were generated with the reference enhancer (hs1617) and the DNVs (hs1617.1 and hs1617.2). X-gal-stained embryos show in vivo enhancer activity (blue) in skeletal structures for each region. Embryos are grouped by genotype (tandem = multiple copies, single = single copy). However, no detectable change in enhancer activity was observed with the presence of either DNV.

**Figure S4:**
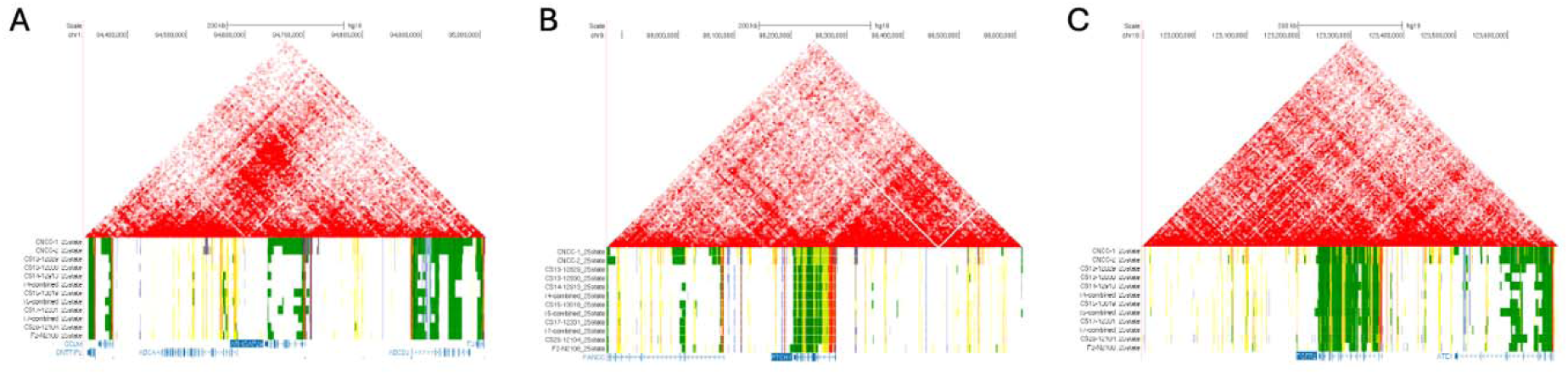
TAD of all DNVs in enhancers. The predicted TAD in cranial neural crest cells for the regions where a DNV was found in a predicted enhancer: ARHGAP29 (**A**), PTCH1 (**B**), and FGFR2 (**C**).

